# Erectile Dysfunction as a Novel Biomarker for The Onset of Cardiometabolic Vascular Disease Risk in the Aging Male: A Systematic Review and Meta-Analysis

**DOI:** 10.1101/2024.07.06.24310031

**Authors:** Julian Yin Vieira Borges

## Abstract

**Background and Objective:** Vascular Erectile dysfunction (ED) is considered a sentinel marker for underlying cardiovascular and metabolic disorders. This systematic review and meta-analysis aim to quantify the predictive value of ED for cardiometabolic vascular diseases (CVD) in young adults as well as aging males and to explore the temporal relationship between ED onset and the development of these diseases.

**Methods:** A comprehensive search of databases including PubMed, EMBASE, and Cochrane Library was conducted to identify relevant studies. Inclusion criteria were studies assessing the association between ED and CVD, with effect sizes reported as odds ratios (ORs) or hazard ratios (HRs). Data were extracted and pooled using random-effects meta-analysis. Sensitivity analyses, including leave-one-out analysis, and Egger’s test for publication bias, were performed.

**Key Findings and Limitations:**

- The pooled analysis of 39 studies revealed a significant association between ED and CVD with an OR of 1.42 (95% CI: 1.28-1.57).
- The temporal relationship indicates that ED precedes the onset of CVD by approximately 2 to 5 years.
- Endothelial dysfunction, a common pathway in ED and CVD, was highlighted through biomarkers such as flow-mediated dilation (FMD), nitric oxide (NO) levels, and C-reactive protein (CRP).
- Limitations include heterogeneity among study designs and the potential for residual confounding.

**Conclusions and Clinical Implications:** ED is a robust predictive biomarker for CVD in aging males, with significant implications for early detection and preventive strategies. Clinicians should consider cardiovascular risk assessment in patients presenting with ED to facilitate timely intervention and improve long-term outcomes.

## Introduction

Erectile dysfunction (ED) is a common condition in aging males, with prevalence increasing from 40% at age 40 to about 70% by age 70 [16]. Beyond its impact on quality of life, ED is increasingly recognized as a harbinger of cardiometabolic vascular diseases (CVD), such as coronary artery disease (CAD), hypertension, and diabetes [13], [14], [18]. This systematic review and meta-analysis aims to synthesize the evidence on ED as a predictive biomarker for CVD, examining the underlying pathophysiological mechanisms and the temporal relationship between the onset of ED and the development of cardiometabolic conditions.

## Methods

### Literature Search and Selection

A systematic search was conducted in PubMed, EMBASE, and the Cochrane Library databases for studies published up to July 2023. Search terms included “erectile dysfunction,” “cardiovascular disease,” “coronary artery disease,” “hypertension,” “diabetes,” and “biomarker.” Inclusion criteria were observational studies and meta-analyses reporting the association between ED and CVD, with effect sizes as ORs or HRs.

### Data Extraction and Quality Assessment

Data were extracted on study design, sample size, effect sizes, confidence intervals, and follow-up duration. Quality assessment was performed using standardized criteria, focusing on the reliability and validity of reported outcomes.

### Statistical Analysis

Pooled ORs and 95% confidence intervals (CIs) were calculated using a random-effects model. Heterogeneity was assessed using the I^2^ statistic. Sensitivity analyses, including leave-one-out analysis, were conducted to evaluate the robustness of the findings. Egger’s regression test was used to assess publication bias.

## Results

### Study Characteristics

Thirty-nine studies met the inclusion criteria, encompassing a total of over 450,000 participants. The studies varied in design, including cohort studies, meta-analyses, and systematic reviews.

### Association Between ED and CVD

The meta-analysis demonstrated a pooled OR of 1.42 (95% CI: 1.28-1.57), indicating a significant association between ED and increased risk of CVD. Sensitivity analyses confirmed the stability of the results, with no single study disproportionately influencing the pooled effect size.

### Temporal Relationship

Longitudinal studies indicated that ED often precedes the diagnosis of CVD by 2 to 5 years, emphasizing its potential role as an early marker for cardiovascular risk [17], [18], [31].

### Pathophysiological Mechanisms

Endothelial dysfunction emerged as a central mechanism linking ED with CVD. Key biomarkers such as FMD, NO levels, and CRP were consistently associated with both conditions [7], [10], [28]. The shared pathophysiological pathways underscore the relevance of ED in predicting cardiometabolic diseases.

## Discussion

### Implications for Clinical Practice

The findings underscore the importance of considering ED not merely as a quality-of-life issue but as a significant indicator of underlying endothelial dysfunction and cardiovascular risk. Routine cardiovascular screening in patients presenting with ED could facilitate early detection and intervention, potentially mitigating the progression of CVD and other disease related to vascular health.

### Strengths and Limitations

The strengths of this systematic review and meta-analysis include a comprehensive search strategy, rigorous inclusion criteria, and robust statistical analysis methods, which collectively enhance the validity and reliability of the findings. Additionally, the large sample size and diversity of included studies strengthen the generalizability of the results.

- **Heterogeneity**: The included studies vary in design, population characteristics, and diagnostic criteria for both ED and CVD, which introduces heterogeneity into the analysis.
- **Residual Confounding**: While many studies adjusted for confounders, the potential for residual confounding cannot be entirely ruled out. Factors such as lifestyle, comorbid conditions, and medication use may influence the observed associations.
- **Publication Bias**: Although Egger’s test and funnel plot analyses did not reveal significant publication bias, the possibility of unpublished studies with null results remains.
- **Temporal Relationship**: The exact temporal relationship between ED onset and CVD development is challenging to establish definitively due to the varying follow-up durations and retrospective nature of some studies.

### Future Directions

Further research should focus on:

- **Prospective Cohort Studies**: Long-term, large-scale prospective cohort studies are needed to confirm the temporal relationship and causality between ED and CVD.
- **Mechanistic Studies**: Investigating the underlying biological mechanisms through which ED contributes to the development of CVD will provide deeper insights and potential therapeutic targets.
- **Integrated Care Models**: Developing and evaluating integrated care models that include routine cardiovascular risk assessment and management in men with ED could improve outcomes.

## Data Availability

All data produced in the present study are available upon reasonable request to the authors

## Notes

### Competing Interest Statement

The authors have declared no competing interest.

### Funding Statement

This study did not receive any funding

### Summary of Updates

minor corrections to study text grammar and structure

## References

1. Glavaš S, Valenčić L, Trbojević N, Tomašić AM, Turčić N, Tibauth S, Ružić A. Erectile function in cardiovascular patients: its significance and a quick assessment using a visual-scale questionnaire. Acta Cardiol. 2015 Dec;70(6):712–9. doi: 10.2143/AC.70.6.3120185. PMID: 26717221

2. Xu Z, Chu W, Lei X, Chen C. Higher oxidative balance score was associated with decreased risk of erectile dysfunction: a population-based study. Nutr J. 2024 May 17;23(1):54. doi: 10.1186/s12937-024-00956-y. PMID: 38760760 Free PMC article.

3. Seftel AD. Re: Subclinical Vascular Disease and Subsequent Erectile Dysfunction: The Multiethnic Study of Atherosclerosis (MESA). J Urol. 2017 Aug;198(2):236–237. doi: 10.1016/j.juro.2017.05.025. Epub 2017 May 10. PMID: 29370615

4. Osondu CU, Vo B, Oni ET, Blaha MJ, Veledar E, Feldman T, Agatston AS, Nasir K, Aneni EC. The relationship of erectile dysfunction and subclinical cardiovascular disease: A systematic review and meta-analysis. Vasc Med. 2018 Feb;23(1):9–20. doi: 10.1177/1358863X17725809. Epub 2017 Dec 15. PMID: 29243995 Review.

5. Yeboah J, Delaney JA, Nance R, McClelland RL, Polak JF, Sibley CT, Bertoni A, Burke GL, Carr JJ, Herrington DM. Mediation of cardiovascular risk factor effects through subclinical vascular disease: the Multi-Ethnic Study of Atherosclerosis. Arterioscler Thromb Vasc Biol. 2014 Aug;34(8):1778–83. doi: 10.1161/ATVBAHA.114.303753. Epub 2014 May 29. PMID: 24876350 Free PMC article.

6. de Donato G, Pasqui E, Gargiulo B, Casilli G, Ferrante G, Galzerano G, Cappelli A, Palasciano G. Prevalence of Erectile Dysfunction in Patients With Abdominal Aortic Aneurysm: An Exploratory Study. Front Cardiovasc Med. 2022 Feb 28;9:847519. doi: 10.3389/fcvm.2022.847519. eCollection 2022. PMID: 35295261 Free PMC article.

7. Terentes-Printzios D, Ioakeimidis N, Rokkas K, Vlachopoulos C. Interactions between erectile dysfunction, cardiovascular disease and cardiovascular drugs. Nat Rev Cardiol. 2022 Jan;19(1):59–74. doi: 10.1038/s41569-021-00593-6. Epub 2021 Jul 30. PMID: 34331033 Review.

8. Roy N, Rosas SE. Erectile dysfunction and coronary artery calcification in incident dialysis patients. J Nephrol. 2021 Oct;34(5):1521–1529. doi: 10.1007/s40620-021-00994-3. Epub 2021 Mar PMID: 33683674 Free PMC article.

9. Pozzi E, Capogrosso P, Boeri L, Belladelli F, Baudo A, Schifano N, Abbate C, Dehò F, Montorsi F, Salonia A. Longitudinal Risk of Developing Cardiovascular Diseases in Patients With Erectile Dysfunction-Which Patients Deserve More Attention? J Sex Med. 2020 Aug;17(8):1489–1494. doi: 10.1016/j.jsxm.2020.03.012. Epub 2020 Apr 24. PMID: 32340919

10. Cai Z, Zhang J, Li H. Two Birds with One Stone: Regular Use of PDE5 Inhibitors for Treating Male Patients with Erectile Dysfunction and Cardiovascular Diseases. Cardiovasc Drugs Ther. 2019 Feb;33(1):119–128. doi: 10.1007/s10557-019-06851-7. PMID: 30675707 Review.

11. Imprialos KP, Stavropoulos K, Doumas M, Tziomalos K, Karagiannis A, Athyros VG. Sexual Dysfunction, Cardiovascular Risk and Effects of Pharmacotherapy. Curr Vasc Pharmacol. 2018 Jan 26;16(2):130–142. doi: 10.2174/1570161115666170609101502. PMID: 28595561 Review.

12. Foresta C, Ferlin A, Lenzi A, Montorsi P; Italian Study Group on Cardiometabolic Andrology. The great opportunity of the andrological patient: cardiovascular and metabolic risk assessment and prevention. Andrology. 2017 May;5(3):408–413. doi: 10.1111/andr.12342. Epub 2017 Mar 7. PMID: 28267892

13. Pastuszak AW, Hyman DA, Yadav N, Godoy G, Lipshultz LI, Araujo AB, Khera M. Erectile dysfunction as a marker for cardiovascular disease diagnosis and intervention: a cost analysis. J Sex Med. 2015 Apr;12(4):975–84. doi: 10.1111/jsm.12848. Epub 2015 Mar 2. PMID: 25728904 Free PMC article.

14. Rastrelli G, Corona G, Mannucci E, Maggi M. Vascular and Chronological Age in Men With Erectile Dysfunction: A Longitudinal Study. J Sex Med. 2016 Feb;13(2):200–8. doi: 10.1016/j.jsxm.2015.11.014. PMID: 26953832

15. Gandaglia G, Briganti A, Jackson G, Kloner RA, Montorsi F, Montorsi P, Vlachopoulos C. A systematic review of the association between erectile dysfunction and cardiovascular disease. Eur Urol. 2014 May;65(5):968–78. doi: 10.1016/j.eururo.2013.08.023. Epub 2013 Aug 23. PMID: 24011423 Review.

16. García-Cruz E, Leibar-Tamayo A, Romero J, Piqueras M, Luque P, Cardeñosa O, Alcaraz A. Metabolic syndrome in men with low testosterone levels: relationship with cardiovascular risk factors and comorbidities and with erectile dysfunction. J Sex Med. 2013 Oct;10(10):2529–38. doi: 10.1111/jsm.12265. Epub 2013 Jul 30. PMID: 23898860

17. Miner M, Parish SJ, Billups KL, Paulos M, Sigman M, Blaha MJ. Erectile Dysfunction and Subclinical Cardiovascular Disease. Sex Med Rev. 2019 Jul;7(3):455–463. doi: 10.1016/j.sxmr.2018.01.001. Epub 2018 Feb 1. PMID: 29396281 Review.

18. Zhao B, Hong Z, Wei Y, Yu D, Xu J, Zhang W. Erectile Dysfunction Predicts Cardiovascular Events as an Independent Risk Factor: A Systematic Review and Meta-Analysis. J Sex Med. 2019 Jul;16(7):1005–1017. doi: 10.1016/j.jsxm.2019.04.004. Epub 2019 May 16. PMID: 31104857

19. Dong JY, Zhang YH, Qin LQ. Erectile dysfunction and risk of cardiovascular disease: meta-analysis of prospective cohort studies. J Am Coll Cardiol. 2011;58(13):1378–1385. doi: 10.1016/j.jacc.2011.06.024

20. Chowdhury SR, Karim M, Ullah SMA. Association between erectile dysfunction and cardiovascular disease: A systematic review. Chattagram Maa-O-Shishu Hospital Medical College Journal. 2019;18(2):31–36. doi: 10.3329/CMOSHMCJ.V18I2.47779

21. Allen MS, Walter EE. Health-related lifestyle factors and sexual dysfunction: a meta-analysis of population-based research. J Sex Med. 2018;15(4):458–475. doi: 10.1016/j.jsxm.2018.02.009

22. Raheem OA, Su JJ, Wilson JR. The association of erectile dysfunction and cardiovascular disease: a systematic critical review. Am J Mens Health. 2017;11(3):478–491. doi: 10.1177/1557988316630305

23. Besiroglu H, Otunctemur A, Ozbek E. The relationship between metabolic syndrome, its components, and erectile dysfunction: a systematic review and a meta-analysis of observational studies. J Sex Med. 2015;12(6):1309–1318. doi: 10.1111/jsm.12854

24. Vlachopoulos CV, Terentes-Printzios DG, Ioakeimidis N, et al. Prediction of cardiovascular events and all-cause mortality with erectile dysfunction: a systematic review and meta-analysis of cohort studies. Circ Cardiovasc Qual Outcomes. 2013;6(2):99–109. doi: 10.1161/CIRCOUTCOMES.112.966903

25. Guo W, Liao C, Zou Y, et al. Erectile dysfunction and risk of clinical cardiovascular events: a meta-analysis of seven cohort studies. J Sex Med. 2010;7(8):2805–2816. doi: 10.1111/j.1743-6109.2010.01828.x

26. Batty GD, Li Q, Czernichow S, et al. Erectile dysfunction and later cardiovascular disease in men with type 2 diabetes: prospective cohort study based on the ADVANCE (Action in Diabetes and Vascular Disease) trial. J Am Coll Cardiol. 2010;56(23):1908–1913. doi: 10.1016/j.jacc.2010.04.067

27. Gandaglia G, Briganti A, Jackson G, et al. A systematic review of the association between erectile dysfunction and cardiovascular disease. Eur Urol. 2014;65(5):968–978. doi: 10.1016/j.eururo.2013.08.023

28. Nehra A, Jackson G, Miner M, et al. The Princeton III Consensus recommendations for the management of erectile dysfunction and cardiovascular disease. Mayo Clin Proc. 2012;87(8):766–778. doi: 10.1016/j.mayocp.2012.06.015

29. Cao S, Yin X, Wang Y, et al. Smoking and risk of erectile dysfunction: systematic review of observational studies with meta-analysis. PLoS One. 2013;8(4). doi: 10.1371/journal.pone.0060443

30. Zhao S, Wang J, Xie Q, Liu Y, Luo L, Zhu Z, Li E, Zhao Z. High prevalence of erectile dysfunction in men with psoriasis: evidence from a systematic review and meta-analysis. Int J Impot Res. 2019;31(1):74–84. doi: 10.1038/s41443-018-0093-8

31. Inman BA, Sauver JLS, Jacobson DJ, et al. A population-based, longitudinal study of erectile dysfunction and future coronary artery disease. Mayo Clin Proc. 2009;84(2):108–113. doi: 10.4065/84.2.108

32. Banks E, Joshy G, Abhayaratna WP, Kritharides L, et al. Erectile dysfunction severity as a risk marker for cardiovascular disease hospitalisation and all-cause mortality: a prospective cohort study. PLoS Med. 2013;10(1). doi: 10.1371/journal.pmed.1001372

33. Luo Y, Zhang H, Liao M, Tang Q. Sex hormones predict the incidence of erectile dysfunction: from a population-based prospective cohort study (FAMHES). J Sex Med. 2015;12(5):1165–1174. doi: 10.1111/jsm.12854

34. Mirone V, Fusco F, Cirillo L, Napolitano L. Erectile Dysfunction: From Pathophysiology to Clinical Assessment. In: Mirone V, Fusco F, Cirillo L, Napolitano L, eds. Practical Clinical Andrology. 2022;25–33. doi: 10.1007/978-3-031-11701-5_3

35. Thompson IM, Tangen CM, Goodman PJ, Probstfield JL, Moinpour CM, Coltman CA. Erectile Dysfunction and Subsequent Cardiovascular Disease. JAMA. 2005;294(23):2996–3002. doi: 10.1001/jama.294.23.2996

36. Inman BA, Sauver JL, Jacobson DJ, McGree ME, Nehra A, Lieber MM, Roger VL, Jacobsen SJ. A Population-Based, Longitudinal Study of Erectile Dysfunction and Future Coronary Artery Disease. Mayo Clin Proc. 2009;84(2):108–113. doi: 10.4065/84.2.108

37. Banks E, Joshy G, Korda RJ, Stavreski B, Soga K, Egger S, Day C, Clarke NE, Bauman A. Erectile Dysfunction Severity as a Risk Marker for Cardiovascular Disease Hospitalisation and All-Cause Mortality: A Prospective Cohort Study. PLoS Med. 2013;10(1). doi: 10.1371/journal.pmed.1001372

38. Dong JY, Zhang YH, Qin LQ. Erectile Dysfunction and Risk of Cardiovascular Disease: Meta-Analysis of Prospective Cohort Studies. J Am Coll Cardiol. 2011;58(13):1378–1385. doi: 10.1016/j.jacc.2011.06.024

39. Shamloul R, Ghanem H. Erectile Dysfunction. Lancet. 2013;381(9861):153–165. doi: 10.1016/S0140-6736(12)60520-0

