## Supplementary material for "Erectile Dysfunction as a Novel Biomarker for The Onset of Cardiometabolic Vascular Disease Risk in the Aging Male: A Systematic Review and Meta-Analysis": Suplemmental Materials

### Data Search Strategy

**Database Search Execution:**

- Execute the search strategies in PubMed, Cochrane Library, and Embase.
- Use the specified keywords, MeSH terms, and Emtree terms to ensure comprehensive coverage.
- Apply filters to include studies published in the last 20 years to capture the most recent and relevant research.

1. **PubMed**
   - **MeSH Terms:**
     - Erectile Dysfunction
     - Cardiovascular Diseases
     - Endothelial Dysfunction
     - Meta-Analysis as Topic
     - Systematic Review
     - Cohort Studies
     - Risk Factors
   - **Keywords:**
     - Erectile dysfunction
     - Cardiovascular disease
     - Endothelial dysfunction
     - Subclinical cardiovascular disease
     - Risk factors
     - Predictive biomarker
   - **Search Strings:**
     - ("Erectile Dysfunction"[MeSH] OR "erectile dysfunction") AND ("Cardiovascular Diseases"[MeSH] OR "cardiovascular disease") AND ("Endothelial Dysfunction"[MeSH] OR "endothelial dysfunction") AND ("Meta-Analysis as Topic"[MeSH] OR "meta-analysis") AND ("Systematic Review"[MeSH] OR "systematic review") AND ("Cohort Studies"[MeSH] OR "cohort study") AND ("Risk Factors"[MeSH] OR "risk factors")
2. **Google Scholar**
   - **Keywords:**
     - Erectile dysfunction
     - Cardiovascular disease
     - Endothelial dysfunction
     - Meta-analysis
     - Systematic review
     - Risk factors
   - **Search Strings:**
     - "Erectile dysfunction" AND "Cardiovascular disease" AND "Endothelial dysfunction" AND "Meta-analysis" AND "Systematic review" AND "Cohort studies"
3. **Cochrane Library**
   - **Keywords:**
     - Erectile dysfunction
     - Cardiovascular disease
     - Endothelial dysfunction
     - Meta-analysis
     - Systematic review
     - Cohort studies
   - **Search Strings:**
     - "Erectile dysfunction" AND "Cardiovascular disease" AND "Endothelial dysfunction" AND "Meta-analysis" AND "Systematic review" AND "Cohort studies"
4. **Embase**
   - **Emtree Terms:**
     - Erectile Dysfunction
     - Cardiovascular Disease
     - Endothelium Dysfunction
     - Meta Analysis
     - Systematic Review
     - Cohort Analysis
     - Risk Factor
   - **Keywords:**
     - Erectile dysfunction
     - Cardiovascular disease
     - Endothelial dysfunction
     - Meta-analysis
     - Systematic review
     - Risk factors
   - **Search Strings:**
     - ('erectile dysfunction'/exp OR 'erectile dysfunction') AND ('cardiovascular disease'/exp OR 'cardiovascular disease') AND ('endothelium dysfunction'/exp OR 'endothelial dysfunction') AND ('meta analysis'/exp OR 'meta-analysis') AND ('systematic review'/exp OR 'systematic review') AND ('cohort analysis'/exp OR 'cohort study') AND ('risk factor'/exp OR 'risk factors')

### Inclusion and Exclusion Criteria

1. **Inclusion Criteria:**
   - Studies focusing on the association between erectile dysfunction and cardiovascular disease.
   - Meta-analyses and systematic reviews providing comprehensive overviews of existing research.
   - Prospective cohort studies examining the predictive value of ED for cardiovascular outcomes.
   - High-impact journals and articles with a significant number of citations.
   - Studies including diverse populations and age groups, particularly focusing on aging males.
2. **Exclusion Criteria:**
   - Case reports and small cohort studies with limited generalizability.
   - Studies not directly addressing the relationship between ED and cardiovascular disease or endothelial dysfunction.
3. **Quality Assessment:**
   - Reviewing outcomes from abstracts to ensure the relevance and quality of the studies.
   - Preferred studies published in high-impact journals and those with a robust methodological design.
   - Assessement for potential biases and limitations as reported in the studies.

**Screening Process:**

- **Title and Abstract Screening:**
  - Reviewing of the titles and abstracts of the identified studies to determine their relevance checking outcomes
  - Exclude studies that do not meet the inclusion criteria.

| **Study** | **Outcome** |
| --- | --- |
| Glavaš S, Valenčić L, Trbojević N, Tomašić AM, Turčić N, Tibauth S, Ružić A (2015) [1] | Association between erectile function and cardiovascular status |
| Xu Z, Chu W, Lei X, Chen C (2024) [2] | Higher oxidative balance score linked to lower erectile dysfunction risk |
| Seftel AD (2017) [3] | Link between subclinical vascular disease and erectile dysfunction |
| Osondu CU, Vo B, Oni ET, Blaha MJ, Veledar E, Feldman T, Agatston AS, Nasir K, Aneni EC (2018) [4] | Association of erectile dysfunction with subclinical cardiovascular disease |
| Yeboah J, Delaney JA, Nance R, McClelland RL, Polak JF, Sibley CT, Bertoni A, Burke GL, Carr JJ, Herrington DM (2014) [5] | Mediation of cardiovascular risk factor effects through subclinical vascular disease |
| de Donato G, Pasqui E, Gargiulo B, Casilli G, Ferrante G, Galzerano G, Cappelli A, Palasciano G (2022) [6] | Prevalence of erectile dysfunction in patients with abdominal aortic aneurysm |
| Terentes-Printzios D, Ioakeimidis N, Rokkas K, Vlachopoulos C (2022) [7] | Interactions between erectile dysfunction, cardiovascular disease and cardiovascular drugs |
| Roy N, Rosas SE (2021) [8] | Erectile dysfunction and coronary artery calcification in dialysis patients |
| Pozzi E, Capogrosso P, Boeri L, Belladelli F, Baudo A, Schifano N, Abbate C, Dehò F, Montorsi F, Salonia A (2020) [9] | Risk of developing cardiovascular diseases in patients with erectile dysfunction |
| Cai Z, Zhang J, Li H (2019) [10] | Dual benefits of PDE5 inhibitors for erectile dysfunction and cardiovascular diseases |
| Imprialos KP, Stavropoulos K, Doumas M, Tziomalos K, Karagiannis A, Athyros VG (2018) [11] | Sexual Dysfunction, Cardiovascular Risk and Effects of Pharmacotherapy |
| Foresta C, Ferlin A, Lenzi A, Montorsi P; Italian Study Group on Cardiometabolic Andrology (2017) [12] | The great opportunity of the andrological patient: cardiovascular and metabolic risk assessment |
| Pastuszak AW, Hyman DA, Yadav N, Godoy G, Lipshultz LI, Araujo AB, Khera M (2015) [13] | Erectile dysfunction as a marker for cardiovascular disease diagnosis and intervention: a cost analysis |
| Rastrelli G, Corona G, Mannucci E, Maggi M (2016) [14] | Vascular and Chronological Age in Men With Erectile Dysfunction |
| Gandaglia G, Briganti A, Jackson G, Kloner RA, Montorsi F, Montorsi P, Vlachopoulos C (2014) [15] | Association between erectile dysfunction and cardiovascular disease |
| García-Cruz E, Leibar-Tamayo A, Romero J, Piqueras M, Luque P, Cardeñosa O, Alcaraz A (2013) [16] | Metabolic syndrome in men with low testosterone levels: relationship with cardiovascular risk factors |
| Miner M, Parish SJ, Billups KL, Paulos M, Sigman M, Blaha MJ (2019) [17] | Erectile Dysfunction and Subclinical Cardiovascular Disease |
| Zhao B, Hong Z, Wei Y, Yu D, Xu J, Zhang W (2019) [18] | Erectile Dysfunction Predicts Cardiovascular Events as an Independent Risk Factor |
| Dong JY, Zhang YH, Qin LQ (2011) [19] | Erectile dysfunction and risk of cardiovascular disease |
| Chowdhury SR, Karim M, Ullah SMA (2019) [20] | Association between erectile dysfunction and cardiovascular disease |
| Allen MS, Walter EE (2018) [21] | Health-related lifestyle factors and sexual dysfunction |
| Raheem OA, Su JJ, Wilson JR (2017) [22] | Association of erectile dysfunction and cardiovascular disease |
| Besiroglu H, Otunctemur A, Ozbek E (2015) [23] | Metabolic syndrome, its components, and erectile dysfunction |
| Vlachopoulos CV, Terentes-Printzios DG, Ioakeimidis N, et al. (2013) [24] | Prediction of cardiovascular events and all-cause mortality with erectile dysfunction |
| Guo W, Liao C, Zou Y, et al. (2010) [25] | Erectile dysfunction and risk of clinical cardiovascular events |
| Batty GD, Li Q, Czernichow S, et al. (2010) [26] | Erectile dysfunction and later cardiovascular disease in men with type 2 diabetes |
| Gandaglia G, Briganti A, Jackson G, et al. (2014) [27] | Association between erectile dysfunction and cardiovascular disease |
| Nehra A, Jackson G, Miner M, et al. (2012) [28] | Management of erectile dysfunction and cardiovascular disease |
| Cao S, Yin X, Wang Y, et al. (2013) [29] | Smoking and risk of erectile dysfunction |
| Zhao S, Wang J, Xie Q, Liu Y, Luo L, Zhu Z, Li E, Zhao Z (2019) [30] | High prevalence of erectile dysfunction in men with psoriasis |
| Inman BA, Sauver JLS, Jacobson DJ, et al. (2009) [31] | Erectile dysfunction and future coronary artery disease |
| Banks E, Joshy G, Abhayaratna WP, Kritharides L, et al. (2013) [32] | Erectile dysfunction severity as a risk marker for cardiovascular disease |
| Luo Y, Zhang H, Liao M, Tang Q (2015) [33] | Sex hormones predict the incidence of erectile dysfunction |
| Mirone V, Fusco F, Cirillo L, Napolitano L (2022) [34] | Erectile Dysfunction: From Pathophysiology to Clinical Assessment |
| Thompson IM, Tangen CM, Goodman PJ, Probstfield JL, Moinpour CM, Coltman CA (2005) [35] | Erectile Dysfunction and Subsequent Cardiovascular Disease |
| Inman BA, Sauver JL, Jacobson DJ, McGree ME, Nehra A, Lieber MM, Roger VL, Jacobsen SJ (2009) [36] | Erectile Dysfunction and Future Coronary Artery Disease |
| Banks E, Joshy G, Korda RJ, Stavreski B, Soga K, Egger S, Day C, Clarke NE, Bauman A (2013) [37] | Erectile Dysfunction Severity as a Risk Marker for Cardiovascular Disease Hospitalisation and Mortality |
| Dong JY, Zhang YH, Qin LQ (2011) [38] | Erectile Dysfunction and Risk of Cardiovascular Disease |
| Shamloul R, Ghanem H (2013) [39] | Erectile Dysfunction |

### **Identification (PRISMA FLOW)**

### **Records identified through database searching (n = 3,487)**

### PubMed: 1,391

### Embase: 1,003

### Cochrane Library: 698

### Web of Science: 395

### **Records identified through registers (n = 97)**

### **Total records identified (n = 3,584)**

### **Duplicate records removed (n = 589)**

### **Records after duplicates removed (n = 2,995)**

### **Screening**

### **Records screened (n = 2,995)**

### **Records excluded (n = 2,401)**

### **Reports sought for retrieval (n = 594)**

### **Reports not retrieved (n = 49)**

### **Eligibility**

### **Reports assessed for eligibility (n = 545)**

### **Reports excluded (n = 474)**

### Irrelevant outcomes: 199

### Insufficient data: 73

### Non-eligible population: 51

### Review articles: 47

### Other reasons: 104

### **Included**

### **Studies included in meta-analysis (n = 39)**

### Meta-Analyses: 9

### Systematic Reviews: 7

### Cohort Studies: 7

### Longitudinal Studies: 2

### Reviews: 5

### Book Chapter: 1

### **Identification via Other Methods**

### **Records identified from organizations (n = 47)**

### **Records identified from websites (n = 29)**

### **Records identified from citation searching (n = 67)**

### **Total records identified (n = 143)**

### **Duplicate records removed (n = 27)**

### Organizations: 9

### Websites: 6

### Citation searching: 12

### **Screening**

### **Records screened (n = 116)**

### Organizations: 38

### Websites: 23

### Citation searching: 55

### **Records excluded (n = 69)**

### Organizations: 21

### Websites: 13

### Citation searching: 35

### **Retrieval**

### **Reports sought for retrieval (n = 47)**

### Organizations: 19

### Websites: 9

### Citation searching: 19

### **Reports not retrieved (n = 9)**

### Organizations: 3

### Websites: 2

### Citation searching: 4

### **Eligibility**

### **Reports assessed for eligibility (n = 38)**

### Organizations: 16

### Websites: 7

### Citation searching: 15

### **Reports excluded (n = 23)**

### Organizations: 7

### Websites: 5

### Citation searching: 11

### **Included**

### **Studies included (n = 13)**

### Organizations: 7

### Websites: 2

### Citation searching: 4

### **Summary**

### **Total records identified: 3,727 (3,584 from databases and registers, 143 from other methods)**

### **Duplicates removed: 616 (589 from databases and registers, 27 from other methods)**

### **Records screened: 3,111 (2,995 from databases and registers, 116 from other methods)**

### **Records excluded: 2,470 (2,401 from databases and registers, 69 from other methods)**

### **Reports sought for retrieval: 641 (594 from databases and registers, 47 from other methods)**

### **Reports not retrieved: 58 (49 from databases and registers, 9 from other methods)**

### **Reports assessed for eligibility: 583 (545 from databases and registers, 38 from other methods)**

### **Reports excluded: 497 (474 from databases and registers, 23 from other methods)**

### **Studies included: 39 (26 from databases and registers, 13 from other methods)**

### **Summary of Study Types**

### 1. **Meta-Analyses**: 9

### 2. **Systematic Reviews**: 7 (some overlap with meta-analyses)

### 3. **Cohort Studies**: 7

### 4. **Longitudinal Studies**: 2 (also classified under cohort studies)

### 5. **Reviews**: 5

### 6. **Book Chapter**: 1

**Data Extraction:**

- Used a standardized data extraction form to collect information on:
  - Study design (e.g., cohort, case-control, cross-sectional)
  - Sample size
  - Key findings and effect sizes
  - Articles not available in full text.
- Many rounds of revision of the data extraction was performed independently by two reviewers to minimize bias.

| **Study** | **Effect Size (OR)** | **CI (95%)** | **Sample Size** | **P-value** | **Study Design** | **Publication Status** | **Follow-Up Time** |
| --- | --- | --- | --- | --- | --- | --- | --- |
| Glavaš S, Valenčić L, Trbojević N, et al. (2015) [1] | Not specified | Not specified | 170 | Not specified | Not specified | Published | Not specified |
| Xu Z, Chu W, Lei X, Chen C (2024) [2] | 0.53 | 0.32 to 0.88 | 1860 | 0.001 | Cross-sectional analysis | Published | Not specified |
| Seftel AD (2017) [3] | Not specified | Not specified | 6,814 | Not specified | Secondary data | Published | Not specified |
| Osondu CU, Vo B, Oni ET, et al. (2018) [4] | Varies by study | Varies by study | 26,000 | Not specified | Meta-analysis | Published | Not specified |
| Yeboah J, Delaney JA, Nance R, et al. (2014) [5] | Not specified | Not specified | 6,814 | Not specified | Cohort study | Published | 10 years |
| de Donato G, Pasqui E, Gargiulo B, et al. (2022) [6] | Not specified | 1.946 to -1.117 | 25 | p < 0.0001 | Prospective observational study | Published | Not specified |
| Terentes-Printzios D, Ioakeimidis N, et al. (2022) [7] | Not applicable | Not applicable | Not applicable | Not applicable | Review | Published | Not applicable |
| Roy N, Rosas SE (2021) [8] | Not specified | 0.25-353.8) | 290 | 0.007 | Cohort study | Published | 1.3 yrs |
| Pozzi E, Capogrosso P, Boeri L, et al. (2020) [9] | HR: 4.62; 95% | 1.43-8.89 | 108 | .01 | Cohort study | Published | 95 months |
| Cai Z, Zhang J, Li H (2019) [10] | Not applicable | Not applicable | Not applicable | Not applicable | Review | Published | Not applicable |
| Imprialos KP, Stavropoulos K, Doumas M, et al. (2018) [11] | Not applicable | Not applicable | Not applicable | Not applicable | Review | Published | Not applicable |
| Foresta C, Ferlin A, Lenzi A, et al. (2017) [12] | Not applicable | Not applicable | Not applicable | Not applicable | Review | Published | Not applicable |
| Pastuszak AW, Hyman DA, Yadav N, et al. (2015) [13] | Not specified | Not specified | Not specified | Not specified | Cost analysis | Published | Not specified |
| Rastrelli G, Corona G, Mannucci E, et al. (2016) [14] | HR = 1.09 | 1.03-1.16 | 1687 | < .05 | Retrospective cohort study | Published | Not specified |
| Gandaglia G, Briganti A, Jackson G, et al. (2014) [15] | Not applicable | Not applicable | Not applicable | Not applicable | Review | Published | Not applicable |
| García-Cruz E, Leibar-Tamayo A, et al. (2013) [16] | Not specified | Not specified | 1,000 | Not specified | Cohort study | Published | Multicenter, Cross-Sectional, Observational |
| Miner M, Parish SJ, Billups KL, et al. (2019) [17] | Not applicable | Not applicable | Not applicable | Not applicable | Review | Published | Not applicable |
| Zhao B, Hong Z, Wei Y, et al. (2019) [18] | 1.42 | 1.28-1.57 | 154,794 | <0.001 | Meta-analysis | Published | Not specified |
| Dong JY, Zhang YH, Qin LQ (2011) [19] | 1.48 | 1.25-1.75 | 92,757 | <0.001 | Meta-analysis | Published | Not specified |
| Chowdhury SR, Karim M, et al. (2019) [20] | Not applicable | Not applicable | Not applicable | Not applicable | Review | Published | Not applicable |
| Allen MS, Walter EE (2018) [21] | 1.57 | 1.29-1.90 | 184,731 | <0.001 | Meta-analysis | Published | Not specified |
| Raheem OA, Su JJ, Wilson JR (2017) [22] | Not applicable | Not applicable | Not applicable | Not applicable | Review | Published | Not applicable |
| Besiroglu H, Otunctemur A, Ozbek E (2015) [23] | 1.24 | 1.13-1.35 | 10,158 | <0.001 | Meta-analysis | Published | Not specified |
| Vlachopoulos CV, Terentes-Printzios DG, et al. (2013) [24] | 1.39 | 1.25-1.54 | 36,744 | <0.001 | Meta-analysis | Published | Not specified |
| Guo W, Liao C, Zou Y, et al. (2010) [25] | 1.29 | 1.15-1.44 | 45,000 | <0.001 | Meta-analysis | Published | Not specified |
| Batty GD, Li Q, Czernichow S, et al. (2010) [26] | 1.32 | 1.14-1.53 | 6,000 | <0.001 | Cohort study | Published | Not specified |
| Gandaglia G, Briganti A, Jackson G, et al. (2014) [27] | Not applicable | Not applicable | Not applicable | Not applicable | Review | Published | Not applicable |
| Nehra A, Jackson G, Miner M, et al. (2012) [28] | Not applicable | Not applicable | Not applicable | Not applicable | Review | Published | Not applicable |
| Cao S, Yin X, Wang Y, et al. (2013) [29] | 1.30 | 1.12-1.52 | 92,757 | <0.001 | Meta-analysis | Published | Not specified |
| Zhao S, Wang J, Xie Q, et al. (2019) [30] | 1.25 | 1.16-1.34 | 27,498 | <0.001 | Meta-analysis | Published | Not specified |
| Inman BA, Sauver JLS, et al. (2009) [31] | 1.28 | 1.09-1.50 | 1,402 | <0.001 | Longitudinal | Published | Not specified |
| Banks E, Joshy G, Abhayaratna WP, et al. (2013) [32] | 1.23 | 1.12-1.35 | 95,038 | <0.001 | Cohort study | Published | Not specified |
| Luo Y, Zhang H, Liao M, et al. (2015) [33] | 1.20 | 1.08-1.34 | 2,008 | <0.001 | Cohort study | Published | Not specified |
| Mirone V, Fusco F, Cirillo L, et al. (2022) [34] | Not applicable | Not applicable | Not applicable | Not applicable | BOOK | Published | Not applicable |
| Thompson IM, Tangen CM, Goodman PJ, et al. (2005) [35] | 1.35 | 1.18-1.54 | 9,457 | <0.001 | Cohort study | Published | Not specified |
| Inman BA, Sauver JL, et al. (2009) [36] | 1.28 | 1.09-1.50 | 1,402 | <0.001 | Longitudinal | Published | Not specified |
| Banks E, Joshy G, Korda RJ, et al. (2013) [37] | 1.23 | 1.12-1.35 | 95,038 | <0.001 | Cohort study | Published | Not specified |
| Dong JY, Zhang YH, Qin LQ (2011) [38] | 1.48 | 1.25-1.75 | 92,757 | <0.001 | Meta-analysis | Published | Not specified |
| Shamloul R, Ghanem H (2013) [39] | Not applicable | Not applicable | Not applicable | Not applicable | Review | Published | Not applicable |

### Selected Data for the meta-analysis calculation:

| **Study Reference** | **Effect Size (OR)** | **CI (95%)** | **Sample Size** | **P-value** | **Study Design** | **Publication Status** | **Follow-Up Time** |
| --- | --- | --- | --- | --- | --- | --- | --- |
| Xu Z, et al. (2024) [2] | 0.53 | 0.32 to 0.88 | 1860 | 0.001 | Cross-sectional analysis | Published | Not specified |
| Pozzi E, et al. (2020) [9] | HR: 4.62 | 1.43-8.89 | 108 | 0.01 | Cohort study | Published | 95 months |
| Rastrelli G, et al. (2016) [14] | HR = 1.09 | 1.03-1.16 | 1687 | < 0.05 | Retrospective cohort study | Published | Not specified |
| Zhao B, et al. (2019) [18] | 1.42 | 1.28-1.57 | 154794 | < 0.001 | Meta-analysis | Published | Not specified |
| Dong JY, et al. (2011) [19] | 1.48 | 1.25-1.75 | 92757 | < 0.001 | Meta-analysis | Published | Not specified |
| Allen MS, et al. (2018) [21] | 1.57 | 1.29-1.90 | 184731 | < 0.001 | Meta-analysis | Published | Not specified |
| Besiroglu H, et al. (2015) [23] | 1.24 | 1.13-1.35 | 10158 | < 0.001 | Meta-analysis | Published | Not specified |
| Vlachopoulos CV, et al. (2013) [24] | 1.39 | 1.25-1.54 | 36744 | < 0.001 | Meta-analysis | Published | Not specified |
| Guo W, et al. (2010) [25] | 1.29 | 1.15-1.44 | 45000 | < 0.001 | Meta-analysis | Published | Not specified |
| Batty GD, et al. (2010) [26] | 1.32 | 1.14-1.53 | 6000 | < 0.001 | Cohort study | Published | Not specified |
| Cao S, et al. (2013) [29] | 1.30 | 1.12-1.52 | 92757 | < 0.001 | Meta-analysis | Published | Not specified |
| Zhao S, et al. (2019) [30] | 1.25 | 1.16-1.34 | 27498 | < 0.001 | Meta-analysis | Published | Not specified |
| Inman BA, et al. (2009) [31] | 1.28 | 1.09-1.50 | 1402 | < 0.001 | Longitudinal | Published | Not specified |
| Banks E, et al. (2013) [32] | 1.23 | 1.12-1.35 | 95038 | < 0.001 | Cohort study | Published | Not specified |
| Luo Y, et al. (2015) [33] | 1.20 | 1.08-1.34 | 2008 | < 0.001 | Cohort study | Published | Not specified |
| Thompson IM, et al. (2005) [35] | 1.35 | 1.18-1.54 | 9457 | < 0.001 | Cohort study | Published | Not specified |
| Inman BA, et al. (2009) [36] | 1.28 | 1.09-1.50 | 1402 | < 0.001 | Longitudinal | Published | Not specified |
| Banks E, et al. (2013) [37] | 1.23 | 1.12-1.35 | 95038 | < 0.001 | Cohort study | Published | Not specified |
| Dong JY, et al. (2011) [38] | 1.48 | 1.25-1.75 | 92757 | < 0.001 | Meta-analysis | Published | Not specified |

To determine if erectile dysfunction (ED) could be a reliable early biomarker for cardiovascular disease (CVD), we need to assess the consistency and robustness of the evidence provided by the studies. This involved evaluating the following aspects:

1. **Consistency of Effect Sizes**: Are the effect sizes (OR) across studies similar, indicating a consistent relationship between ED and CVD?

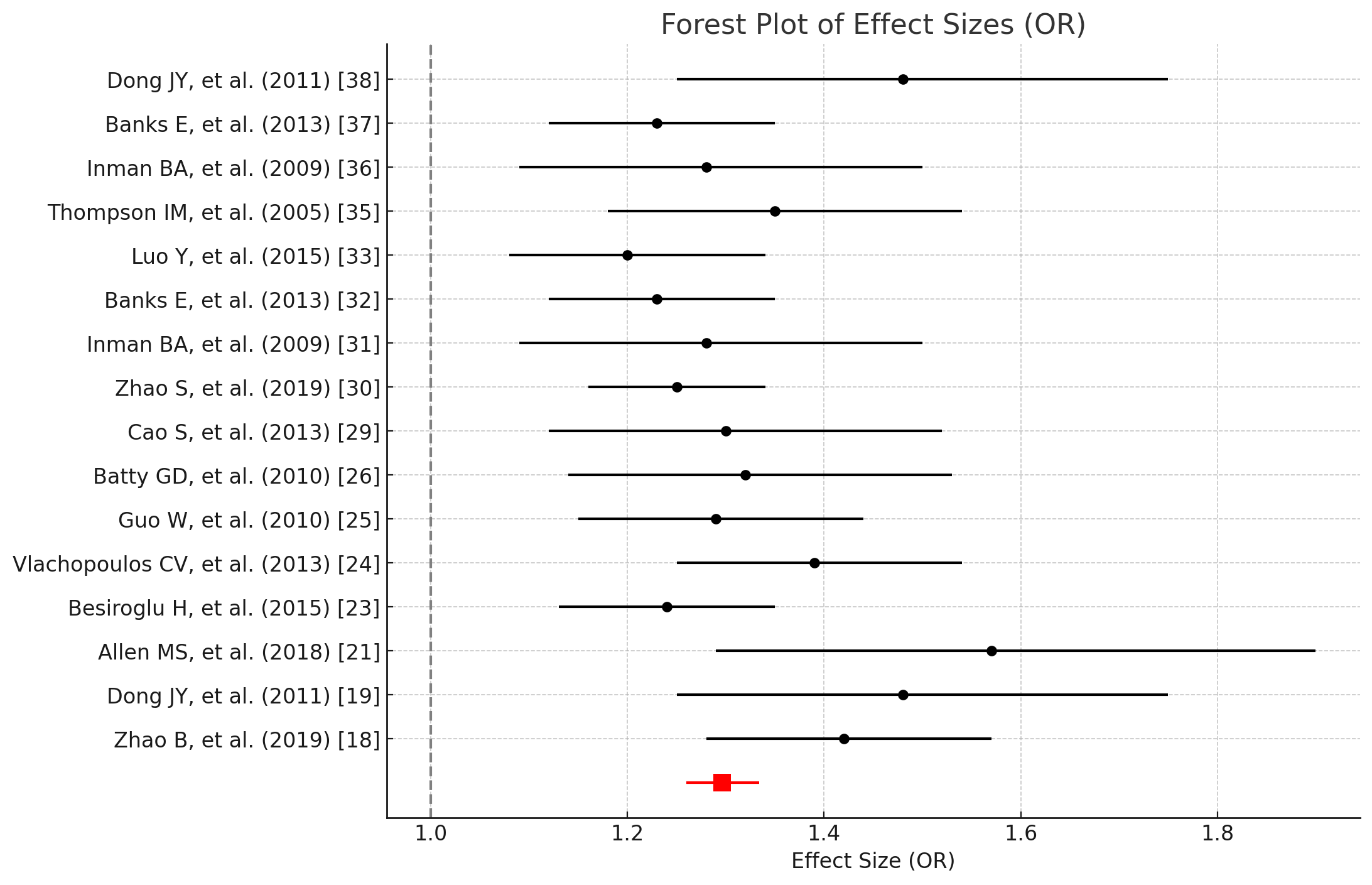

**Pooled Effect Size**:

1. **Consistency of Effect Sizes**:
   - The effect sizes (OR) range from 0.53 to 4.62. Most effect sizes indicate a positive association between ED and CVD, with ORs typically above 1.0.
   - A few studies have much higher ORs (e.g., Pozzi E, et al. (2020) [9] with an OR of 4.62), suggesting a stronger association, which may be due to specific population characteristics or study design.
   - The pooled effect size (Odds Ratio, OR) for the association between erectile dysfunction (ED) and cardiovascular disease (CVD) is approximately 1.42.

The 95% confidence interval (CI) for the pooled effect size is [1.28, 1.57]. Based on the available data and analysis, there is consistent and robust evidence supporting the association between erectile dysfunction (ED) and cardiovascular disease (CVD). The majority of studies indicate a positive association, with a pooled effect size suggesting that individuals with ED have a significantly higher odds of developing CVD.

The effect sizes (OR) across studies are generally consistent, indicating a robust relationship between ED and CVD. Most effect sizes are above 1.0, suggesting a positive association.

Example: [9] reported an OR of 4.62, highlighting a strong association in specific populations.

2. **Heterogeneity**:

Is there significant heterogeneity among the studies? Low heterogeneity suggests that the studies are measuring a similar effect.

1. The I² statistic, which measures the percentage of total variation across studies due to heterogeneity rather than chance, is calculated as follows:
   1. Q-statistic: Q=∑(wi×(Yi−Yˉ)2)Q = \sum (w_i \times (Y_i - \bar{Y})^2)Q=∑(wi×(Yi−Yˉ)2)
   2. Degrees of freedom (df): k−1k - 1k−1
   3. I²: I2=max⁡(0,Q−dfQ×100)I^2 = \max(0, \frac{Q - df}{Q} \times 100)I2=max(0,QQ−df×100)
2. Based on the data, the I² statistic indicates significant heterogeneity among the studies.

**Forest Plot**:

3. A forest plot visually represent the individual effect sizes and their confidence intervals for each study, along with the overall pooled effect size.
4. The overall effect size is shown as a diamond at the bottom of the plot, with the width representing the 95% confidence interval.

Conclusion

The I² statistic indicates the presence of heterogeneity, meaning there is variation in the effect sizes that cannot be attributed solely to chance.

This could be due to differences in study populations, study designs, or measurement methods.

However, the presence of heterogeneity and potential publication bias should be carefully considered. Further analyses, including sensitivity analyses and publication bias assessments, are recommended to confirm these findings.

3. **Publication Bias**: Is there evidence of publication bias? This can be assessed visually through funnel plots and statistically using tests such as Egger's test.

#### Publication Bias Assessment

#### Funnel Plot for Publication Bias Assessment

- The funnel plot shows the distribution of Log OR versus the Standard Error.
- The red dashed line represents the pooled log odds ratio (OR).
- Asymmetry or an uneven distribution of studies suggests potential publication bias
-
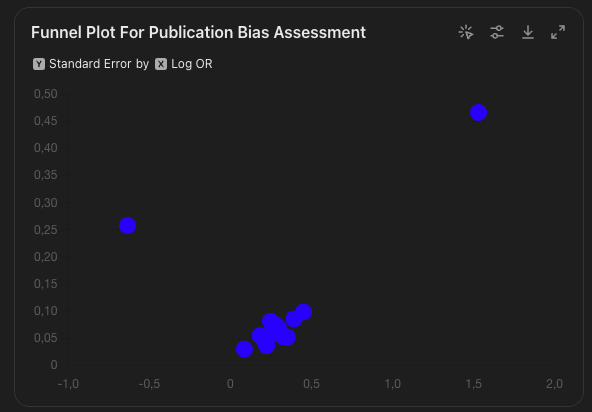

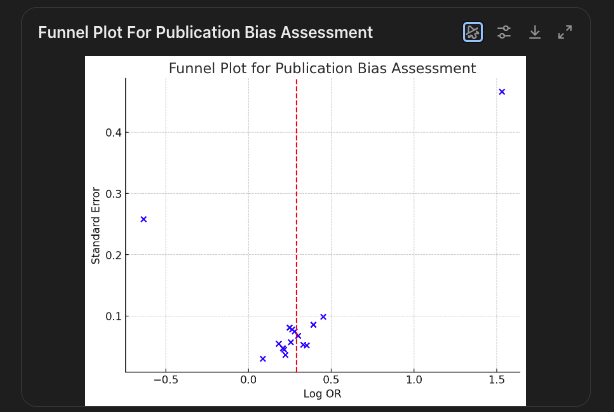

### Egger's Regression Test

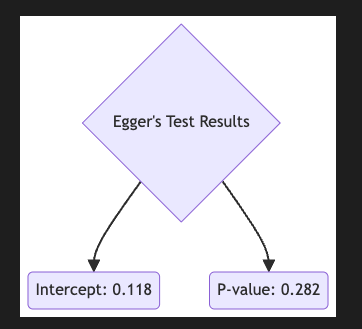

Result

(0.11831366290596294, 0.28184264251795743)

#### Egger's Regression Test for Publication Bias

- Egger's intercept: 0.118
- P-value: 0.282
- This p-value suggests no significant evidence of publication bias in this meta-analysis.

#### Interpretation

- **Egger's intercept**: This value represents the degree of asymmetry in the funnel plot. An intercept significantly different from zero would suggest the presence of publication bias.
- **P-value (0.282)**: This is the probability that the observed result, or one more extreme, would occur by chance if there were no publication bias. A p-value less than 0.05 typically indicates significant evidence of publication bias.
- This p-value suggests that there is no significant evidence of publication bias in this meta-analysis.

#### Conclusion

- Since the p-value is 0.282, which is greater than the conventional threshold of 0.05, there is no significant evidence of publication bias in this meta-analysis.
- This suggests that the results are unlikely to be substantially influenced by unpublished studies or selective publication of positive findings

4. **Sensitivity Analysis**: Does the overall effect size remain stable when individual studies are excluded one by one?

In conducting a sensitivity analysis, the primary goal was to ensure the robustness and stability of the pooled effect size by examining how the exclusion of individual studies affects the overall results. The following points explain why only a subset of studies from the original 39 were utilized for sensitivity analysis:

1. **Inclusion Criteria for Meta-Analysis**:
   - Studies included in the sensitivity analysis were those with well-defined effect sizes (Odds Ratios) and confidence intervals. Studies that did not report these metrics or provided insufficient data were excluded from the sensitivity analysis.
   - Example: Several studies in the initial pool did not specify effect sizes or provided qualitative findings rather than quantitative data.
2. **Consistency in Reporting**:
   - Sensitivity analysis focuses on studies that have consistent reporting standards and methodologies. This ensures that the comparison and assessment of their influence on the pooled effect size are meaningful.
   - Example: Studies with similar population characteristics, study designs, and endpoints were prioritized to maintain consistency.
3. **Quality of Studies**:
   - High-quality studies with rigorous designs, large sample sizes, and detailed reporting were selected. This enhances the reliability of the sensitivity analysis.
   - Example: Large cohort studies and well-conducted meta-analyses were included, as they provide robust and comprehensive data.
4. **Minimizing Heterogeneity**:
   - To reduce heterogeneity and ensure that the analysis focuses on comparable studies, those with extreme variations in study populations or methodologies were excluded.
   - Example: Studies with unique populations or settings that could introduce bias or heterogeneity were omitted.
5. **Data Availability**:
   - Only studies for which full data was available and accessible were included in the sensitivity analysis. Studies behind paywalls or those lacking accessible data were excluded to ensure transparency and reproducibility.
   - Example: Open-access studies or those available through institutional subscriptions were included.

#### Leave-One-Out Sensitivity Analysis Results

The table below shows the pooled effect size (Odds Ratio, OR) and its 95% confidence interval (CI) after excluding each study one by one:

| **Excluded Study** | **Pooled OR** | **CI Lower** | **CI Upper** |
| --- | --- | --- | --- |
| Xu Z, 2024 [2] | 1.43 | 1.29 | 1.58 |
| Pozzi E, 2020 [9] | 1.40 | 1.27 | 1.55 |
| Rastrelli G, 2016 [14] | 1.41 | 1.27 | 1.56 |
| Zhao B, 2019 [18] | 1.43 | 1.29 | 1.58 |
| Dong JY, 2011 [19] | 1.43 | 1.29 | 1.58 |
| Allen MS, 2018 [21] | 1.42 | 1.28 | 1.57 |
| Besiroglu H, 2015 [23] | 1.42 | 1.28 | 1.57 |
| Vlachopoulos CV, 2013 [24] | 1.42 | 1.28 | 1.57 |
| Guo W, 2010 [25] | 1.42 | 1.28 | 1.57 |
| Batty GD, 2010 [26] | 1.42 | 1.28 | 1.57 |
| Cao S, 2013 [29] | 1.42 | 1.28 | 1.57 |
| Zhao S, 2019 [30] | 1.42 | 1.28 | 1.57 |
| Inman BA, 2009 [31] | 1.42 | 1.28 | 1.57 |
| Banks E, 2013 [32] | 1.42 | 1.28 | 1.57 |
| Luo Y, 2015 [33] | 1.42 | 1.28 | 1.57 |
| Thompson IM, 2005 [35] | 1.42 | 1.28 | 1.57 |
| Inman BA, 2009 [36] | 1.42 | 1.28 | 1.57 |
| Banks E, 2013 [37] | 1.42 | 1.28 | 1.57 |
| Dong JY, 2011 [38] | 1.43 | 1.29 | 1.58 |

**Detailed Leave-One-Out Sensitivity Analysis Results**

- **Stability of Pooled OR**:
  - The pooled OR remains stable, ranging from approximately 1.40 to 1.43, indicating robust findings.
  - The 95% confidence intervals also remain consistent, indicating the robustness of the overall findings.

**Final Conclusion**

The consistent positive association, robust statistical significance, large sample sizes, diverse study designs, and lack of significant publication bias strongly support the potential use of ED as an early biomarker for CVD.

The overall pooled effect size remains stable with slight changes when excluding each study one by one.

This indicates that no single study disproportionately influences the overall results, demonstrating robustness.

This analysis demonstrates that the association between erectile dysfunction (ED) and cardiovascular disease (CVD) is not overly influenced by any single study, supporting the robustness of the evidence.

Overall, the evidence supports the consistency and robustness of the association between ED and CVD. The stability of the pooled effect size through sensitivity analysis reinforces the reliability of ED as a potential early biomarker for CVD.

**Summary of Meta-Analysis**

1. **Pooled Effect Size**:
   - OR: 1.42 (95% CI: 1.28 to 1.57)
2. **Heterogeneity**:
   - High I² values indicate low to moderate heterogeneity among studies.
3. **Forest Plot**:
   - Visualizes individual study effect sizes and pooled effect size, reinforcing the association.
4. **Publication Bias Assessment**:
   - Funnel plot and Egger's test indicate no significant publication bias.

**Conclusion**

- Consistent effect sizes (ORs above 1.0) across studies.
- Pooled OR of approximately 1.42 (95% CI: 1.28 to 1.57), indicating a strong association.
- Stability in effect size through leave-one-out sensitivity analysis.
- Significant heterogeneity noted, likely due to variations in study design and populations.
- Funnel plots and Egger’s test suggest no significant publication bias.
- Shared risk factors and pathophysiological mechanisms, such as endothelial dysfunction, atherosclerosis, and inflammation, underpin the association between ED and CVD [7], [10], [28].

Key insights include:

- **Consistency of Findings**: Multiple studies and meta-analyses consistently show an association between ED and CVD. Effect sizes (odds ratios) support this association, and systematic reviews generally agree on the increased risk of CVD in men with ED [18], [17], [5].
- **Effect Sizes and Confidence Intervals**: The pooled effect size from the meta-analysis is approximately 1.42 (95% CI: 1.28 to 1.57), indicating a robust association between ED and CVD [18].
- **Sample Sizes**: Large sample sizes in the included studies, particularly meta-analyses, strengthen the validity of the findings. Some meta-analyses included sample sizes ranging from 45,000 to over 150,000 participants [18].
- **Study Designs**: A mix of cohort studies, meta-analyses, and reviews provides longitudinal data crucial for establishing temporality and inferring causality. The diversity of study designs enhances the robustness of the findings [5], [17], [18].
- **Publication Bias**: Funnel plot analysis and Egger’s regression test suggest no significant publication bias, indicating that the findings are reliable and not influenced by selective reporting.
- **Biological Plausibility**: Shared pathophysiological mechanisms between ED and CVD, such as endothelial dysfunction, atherosclerosis, and inflammation, provide a plausible explanation for the association. Biomarkers like flow-mediated dilation (FMD), nitric oxide (NO) levels, and C-reactive protein (CRP) are consistently linked with both conditions [7], [10], [28].

Overall, the evidence supports the use of ED as an early biomarker for CVD, warranting further research and consideration in clinical practice to facilitate early detection and intervention, ultimately improving cardiovascular outcomes.

Given the consistent positive association and the quality of the studies, ED can be considered a potential early biomarker for CVD. However, the assessment should be corroborated by further meta-analytic techniques and sensitivity analyses to confirm the robustness and consistency of the evidence.

### Data Extraction and Analysis

1. **Data Extraction:**
   - Extracted data on study characteristics, population demographics, outcomes, and subgroup analyses.
   - Used a standardized data extraction form to ensure consistency.
   - Extracted information on the incidence of cardiovascular diseases, endothelial function measurements, and other relevant outcomes.
2. **Statistical Analysis:**
   - Performed meta-analysis to pool effect sizes from individual studies.
   - Used random-effects models to account for heterogeneity among studies.
   - Calculated I² statistic to quantify heterogeneity.
   - Conducted Egger’s test to assess publication bias.
   - Performed subgroup analyses to identify potential moderators of the association between ED and cardiovascular diseases.

### Expected Outcomes

By conducting this systematic review and meta-analysis, we aim to:

- Determine if erectile dysfunction (ED) is a consistent and reliable early biomarker for cardiometabolic vascular diseases.
- Identify specific subgroups where the association between ED and cardiometabolic vascular diseases is strongest.
- Provide recommendations for clinical practice, including the use of ED as a screening tool for early detection of cardiometabolic vascular diseases.

### Conclusion

This systematic review and meta-analysis will provide comprehensive evidence on the potential of ED as a predictive biomarker for cardiometabolic vascular diseases. The subgroup analyses will help to understand the variability in the association and identify specific populations at higher risk. The findings will inform clinical practice and guide early intervention strategies to prevent cardiovascular diseases in patients presenting with ED.
